# Handling onset age inconsistencies in longitudinal healthcare survey data

**DOI:** 10.64898/2026.02.20.26346741

**Authors:** Wanxin Li, Ming Yuan, Yongjin P. Park, Khanh Dao Duc

## Abstract

Longitudinal healthcare surveys frequently contain inconsistencies in self-reported onset ages, where participants report different ages for the same condition between enrollment and follow-up surveys. We propose two methods to handle this challenge. First, we introduce a procedure that aggregates inconsistency patterns to construct participant-level reliability scores, enabling researchers to stratify participants and prioritize analysis on high-reliability cohorts. Second, we present a Bayesian adjustment method that models enrollment and follow-up reports as noisy observations of a latent true onset age, producing adjusted estimates for the inconsistent observations that account for age-dependent and inter-survey-time effects. We evaluate both methods using data from the Canadian Partnership for Tomorrow’s Health. In general, both methods substantially strengthen correlations between biologically related conditions and improve predictive performance across classification and regression tasks. In addition, high-reliability cohorts from reliability score-based stratification reveal more coherent and interpretable disease clustering networks, and Bayesian adjustment shows particularly notable gains when multiple inconsistent variables are adjusted simultaneously. Finally, we provide guidance on choosing between these methods for healthcare practitioners.

**Institutional Review Board (IRB):** The study is approved by the University of British Columbia IRB (IRB #H23-03800).

**Data and Code Availability:** CanPath data are available to researchers through a controlled access process via the CanPath Access Portal (https://portal.canpath.ca). The code is available at https://anonymous.4open.science/r/canpath-FCCF.

## 1 Introduction

Longitudinal healthcare surveys are important for understanding disease etiology and developing predictive models for population health [Caruana et al., 2015, Grant et al., 1995, Maddox and Douglass, 1973]. These surveys collect self-reported information across survey waves, often spanning years or decades. A key variable in many of these surveys is the self-reported onset age, the age at which a participant first experienced or was diagnosed with a particular condition [Benjamin et al., 2017, Veenstra and Syse, 2012], which is important for life-course epidemiology and risk prediction [Skybova et al., 2022].

An onset age inconsistency occurs when a participant reports different onset ages for the same condition across survey waves. It is a type of measurement error as self-reported responses usually suffer from memory lapses, recall bias and careless responding [Shillington et al., 2010]. For example, a participant might report developing diabetes at age 45 during enrollment but report age 52 during follow-up. Studies have shown that such inconsistencies are prevalent across large-scale cohorts [Johnson and Mott, 2001, Parra et al., 2003], yet they pose fundamental challenges: Discarding all inconsistent records can cause substantial data loss, while retaining them introduces measurement error that attenuates effect estimates. While previous work has quantified reliability patterns at the disease level [Johnson and Mott, 2001, Farrer et al., 1989], proposed deterministic reconciliation rules [Andreacchi et al., 2025], and developed structured interview techniques to improve prospective data collection [Axinn and Chardoul, 2021, Caspi et al., 1996], these approaches fail to quantify participant-level reliability or provide statistically principled adjustments that account for age-dependent measurement errors in existing datasets.

In this paper, we propose two methods for handling onset age inconsistencies in longitudinal healthcare survey data. First, we introduce a procedure that aggregates inconsistency patterns across all onset ages to construct participant-level reliability scores. These scores enable researchers to stratify participants by onset age consistency and prioritize high-reliability cohorts or exclude low-reliability cohorts. Second, we present a Bayesian adjustment method that explicitly models measurement error in self-reported onset ages. By treating enrollment and follow-up reports as noisy observations of a latent true onset age, this method produces adjusted estimates for inconsistent observations. We demonstrate both methods using data from the Canadian Partnership for Tomorrow’s Health (CanPath) [Dummer et al., 2018], a cohort of over 97,408 participants followed through enrollment and subsequent surveys, and 57.1% of participants exhibit onset age inconsistencies^2^. We evaluate both methods on association discovery tasks, including correlations between biologically related conditions and disease clustering networks, and on predictive modeling tasks, including classification and regression problems. Finally, we provide practical guidance for choosing between these methods based on sample size constraints, interpretability requirements, and variable characteristics.

## 2 Background and related works

### 2.1 Existing work for handling inconsistent observations in longitudinal healthcare data

Existing approaches for handling inconsistent longitudinal healthcare observations fall into three categories. First, reliability quantification studies measure the consistency of self-reported health information through test-retest assessments and validation against medical records [Johnson and Mott, 2001, Farrer et al., 1989, Louis et al., 2023, Raphael and Marbach, 1997, Dorn et al., 2013, Shillington et al., 2010]. While these studies identify factors affecting reliability (e.g., disease type, recall period), they characterize reliability at the disease level and do not translate these patterns into participant-level metrics. Second, deterministic reconciliation applies rule-based adjudication to resolve contradictions, such as enforcing chronic-condition persistence or prioritizing specific data sources [Andreacchi et al., 2025]. However, these approaches impose fixed solutions without quantifying uncertainty. Third, life history calendars use structured interview techniques to improve recall accuracy through temporal anchoring [Axinn and Chardoul, 2021, Caspi et al., 1996], but require prospective implementation and cannot be applied to existing data.

In the broader statistical literature on longitudinal data, measurement error models treat repeated measures as noisy observations of a latent true value and estimate parameters via likelihood-based methods [Gelman et al., 1995, Quinio and Lam, 2021]. However, these models do not incorporate onset age-specific characteristics, such as age-dependent or inter-survey time effects. Moreover, they lack participant-level interpretability because they do not produce adjusted estimates for individual observations.

Our work addresses these limitations by developing methods that quantify participant-level reliability and provide adjusted onset age estimates that account for age-dependent and inter-survey time effects.

### 2.2 CanPath and its applications

CanPath is a longitudinal healthcare survey study designed to investigate chronic disease prevention across Canada. The entire CanPath cohort contains over 330,000 participants. Our access request, based on variable availability and missingness criteria, yielded data for 97,408 participants. The CanPath study collects comprehensive data, including demographics, lifestyle factors (e.g., physical activity, smoking, alcohol consumption), socioeconomic variables (e.g., education, income), anthropometric measurements (e.g., body mass index), and self-reported disease histories. Participants completed baseline questionnaires at enrollment and were followed through subsequent surveys, typically 5-10 years later. Self-reported onset ages were collected for 55 variables at both enrollment and follow-up. Full names and descriptions of these 55 variables can be found in Appendix A.

Applications of CanPath data span multiple domains of health research. Researchers have utilized the cohort for cancer epidemiology studies, including investigations of breast cancer screening adherence across different provinces [Darvishian et al., 2025], colorectal cancer screening patterns [Darvishian et al., 2023], and lifestyle factors associated with lung cancer risk among never smokers [Murphy et al., 2022]. In chronic disease research, CanPath has enabled studies on cardiovascular disease risk factors, metabolic conditions, and dietary patterns [Shah et al., 2025]. Additionally, CanPath has supported COVID-19 research through national surveillance efforts Nejatinamini et al. [2025]. Linkage with provincial administrative health databases and environmental data from the Canadian Urban Environmental Health Research Consortium [Brook et al., 2018] enables investigation of complex interactions between genetics, lifestyle, environment, and health outcomes [Kuczynski et al., 2024].

## 3 Methods

### 3.1 Reliability score-based stratification

We construct and use participant-level reliability scores using the following steps.

Let **X** ∈ ℝ^*n×m*^ denote the dataset where *n* is the number of participants and *m* is the number of variables, with *X*_*ij*_ denoting participant *i*’s value for variable *j*. Let 𝒜 ⊆ {1, …, *m*} denote the set of onset age variables observed at both enrollment and follow-up, with |𝒜| = *p*. For *j* ∈ 𝒜, let 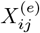 and 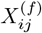 denote participant *i*’s reported onset ages for variable *j* at enrollment and follow-up, respectively.

#### Data preparation

We construct the age difference matrix **D** ∈ ℝ^*n×p*^ with entries:

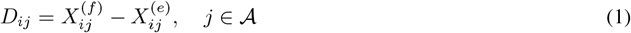

where *D*_*ij*_ is missing if either 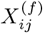 or 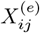 is unavailable. We exclude participants with a missing rate exceeding a threshold *ρ*, yielding *n*^*′*^ participants for the following.

#### Matrix completion

We assume that participant reliability depends only on the magnitude of onset age discrepancies, not their direction (i.e., whether ages are over- or under-reported at follow-up)^3^. Under this assumption, we apply SoftImpute [Mazumder et al., 2010] to impute missing values in **D**^4^, yielding a completed matrix 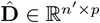, then compute element-wise absolute values 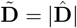.

#### Dimension reduction

We apply principal component analysis (PCA) [Abdi and Williams, 2010] to 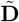 to retain *K* components.

#### Score construction

Let *z*_*ik*_ denote participant *i*’s score on component *k*, and let *w*_*k*_ denote the explained variance ratio of component *k*. We compute the raw reliability score for participant *i* using:

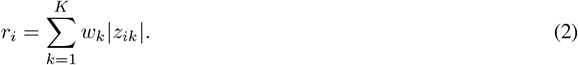

We take absolute values of the component scores because the sign of a PCA component indicates the direction of deviation from the mean, whereas reliability depends only on the magnitude of deviation. Larger values of *r*_*i*_ indicate greater deviations from consistent response patterns.

#### Normalization

To obtain the final reliability scores, we apply quantile normalization [Bolstad et al., 2003] to transform the raw scores to a uniform distribution on [0, 1], with inversion such that higher values indicate greater reliability.

#### Stratification

The reliability scores enable researchers to stratify participants based on response consistencies. When the reliability score distribution exhibits clear multimodality, we can apply k-means clustering [Hartigan and Wong, 1979] to automatically identify thresholds. For unimodal distributions, we recommend examining the distribution’s spread; if narrowly peaked, stratification may be unnecessary. Otherwise, researchers can use percentile-based thresholds (e.g., median) based on the sample size constraints.

### 3.2 Bayesian adjustment

Alternatively, we can directly adjust inconsistent onset age observations by modeling measurement error and computing posterior estimates of the latent values.

For *j* ∈ 𝒜, let 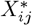 denote participant *i*’s latent true value for variable *j*. Let 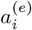 and 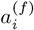 denote participant *i*’s ages at enrolment and follow-up, respectively.

#### Measurement error model

We model the observed onset ages at enrollment 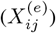 and follow-up 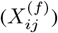 as noisy realizations of a latent true value 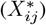:

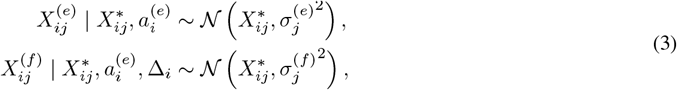

where 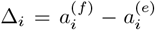, which is the time gap between follow-up and enrollment. We assume the measurement errors are independent conditional on 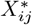 because enrollment and follow-up responses represent separate recall events occurring years apart, and no access to previous responses.

#### Variance parameterization

To encode the assumption that recall accuracy decreases with age and deteriorates further at follow-up [Rhodes et al., 2019], we parameterize the variances as:

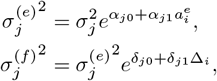

where *α*_*j*1_, *δ*_*j*0_, *δ*_*j*1_ ≥ 0. The constraint *α*_*j*1_ ≥ 0 ensures enrollment variance increases with age, while *δ*_*j*0_, *δ*_*j*1_ ≥ 0 ensures follow-up variance exceeds enrollment variance and increases with longer inter-survey intervals.

#### Parameter estimation

To estimate the variance parameters 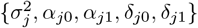 for each onset age variable *j*, we leverage the observed age differences *D*_*ij*_. Let 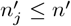 denote the number of participants with observed values for both 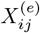 and 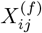. Since 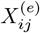 and 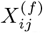 are conditionally independent given 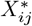 and both normally distributed (see Equation (3)), their difference follows [Cramér, 1999]:

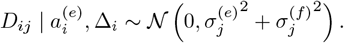

We estimate the parameters by maximizing the log-likelihood:

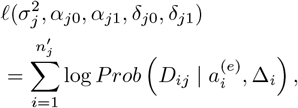

where the sum is over all participants with observed values for both 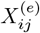 and 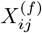.

#### Posterior imputation

Assuming a diffuse Normal prior on 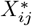, the posterior distribution is:

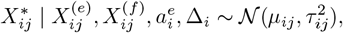

where the posterior mean and variance are:

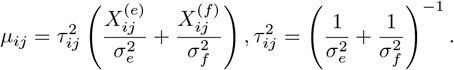

The adjusted value 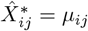 is a precision-weighted average that assigns greater weight to the enrollment observation as it has lower estimated variance. Derivations can be found in Appendix B.

## 4 Experiments and implementation

### 4.1 Reliability score-based stratification

#### Experiments

We evaluated reliability score-based stratification on two downstream tasks: association discovery and predictive modeling. For each task, we stratified participants into high- and low-reliability cohorts based on the median reliability score, excluding participants with missing values on the task-specific variables. For all onset ages mentioned in this section, they refer to the onset ages during enrollment^5^.

In Section 5.2.2, for association discovery, within each of the high- and low-reliability cohorts, we examined pairwise Pearson correlations between onset ages of biologically related conditions and constructed correlation networks to identify disease clusters. The correlation analyses include:

(*co*_11_) Asthma, high cholesterol, high blood pressure, heart attack, stroke onset ages [Garg et al., 2023, Tattersall et al., 2016, Wang et al., 2020, Khoja et al., 2023, Domanski et al., 2020] (mean 458 participants per pairwise analysis);

(*co*_12_) Hearing loss, swimmer’s ear, tinnitus and vertigo onset ages [Wiegand et al., 2019, Azevedo et al., 2022, Ihler et al., 2024, Reisinger et al., 2023, Miura et al., 2017] (mean 39 participants per pairwise analysis).

For network analysis:

(*co*_13_) Correlation networks on 45 disease onset age variables with positive correlations out of 55 onset age variables (mean 151 participants per pairwise analysis).

In Section 5.2.3, for predictive modeling, for each of the high- and low-reliability cohorts, we evaluated classification tasks (predicting disease status) and regression tasks (predicting onset age) using 5-fold cross-validation. For classification, we applied Logistic Regression, Random Forest, Decision Tree, Gradient Boosting, and Support Vector Machine (SVM) with precision and recall as metrics. For regression, we applied Linear Regression, Lasso, Ridge, Random Forest, and SVM with Mean Absolute Error (MAE) and Root Mean Squared Error (RMSE) as metrics. The classification tasks include:

(*cl*_11_) Predicting depression status using sex, age, and anxiety status (2198 participants);

(*cl*_12_) Predicting high blood sugar status using age, sex, high cholesterol status, diabetes status, income, physical activity level, smoking frequency, and alcohol frequency (2953 participants);

(*cl*_13_) Predicting diabetes status using age, sex, education level, income, body mass index, physical activity level, smoking frequency, and alcohol frequency (54 participants).

The regression tasks include:

(*re*_11_) Predicting depression onset age using age, sex, and anxiety onset age (500 participants);

(*re*_12_) Predicting high blood sugar onset age using age, sex, high cholesterol onset age, diabetes onset age, income, physical activity level, smoking frequency, and alcohol frequency (182 participants);

(*re*_13_) Predicting hearing loss onset age using tinnitus onset age, and tinnitus treatment status (1032 participants).

#### Implementation

In data preparation, we set the missing rate threshold *ρ* = 53*/*55 to exclude 74% of participants with fewer than two observed onset age differences across 55 onset age variables, leaving 25,018 participants for analysis. In dimension reduction, we used *K* = 10 principal components, which capture 37% of the total variance in the absolute age difference matrix.

### 4.2 Bayesian adjustment

#### Experiments

We evaluated Bayesian adjustment on association discovery and predictive modeling tasks. In Section 5.3.1, for association discovery, we computed Pearson correlations between pairs of onset age variables using enrollment variables, follow-up variables, and Bayesian-adjusted values (applied to both variables). We selected onset age variable pairs with established positive biological associations:

(*co*_21_) Anxiety and depression [Jacobson and Newman, 2017] (260 participants),

(*co*_22_) High blood pressure and heart attack [Psaty et al., 2001] (474 participants),

(*co*_23_) High cholesterol and high blood pressure [Grundy et al., 2005] (1554 participants).

In Section 5.3.2, for predictive modeling, we evaluated classification and regression tasks with and without Bayesian adjustment using 5-fold cross-validation. For each task, we used the set of models and metrics as in Section 4.1. In addition, we accompanied all metrics with 95% confidence intervals (CIs) computed via non-parametric bootstrapping across the cross-validation folds. Onset age variables with potentially inconsistent enrollment and follow-up observations are marked with ∗; these are the variables to which we applied Bayesian adjustment. The classification tasks include:

(*cl*_21_) Predicting depression status using age, anxiety onset age (∗) (335 participants),

(*cl*_22_) Predicting diabetes status using age, high blood sugar onset age, and high blood pressure onset age (∗) (579 participants).

The regression tasks include:

(*re*_21_) Predicting diabetes onset age using age, sex, high blood pressure onset age (∗), and high cholesterol onset age (∗) (314 participants),

(*re*_22_) Predicting high cholesterol onset age using age, sex, high blood pressure onset age (∗), and high blood sugar status (1380 participants).

#### Implementation

In parameter estimation, to enforce the constraints *α*_*j*1_, *δ*_*j*0_, *δ*_*j*1_ ≥ 0 during maximum likelihood estimation, we reparameterize these parameters using exponential transformations. We maximize the log-likelihood using multiple solvers (L-BFGS-B [Byrd et al., 1995], SLSQP [Kraft, 1988], and TNC [Nash, 1984]) and select the solution with the lowest negative log-likelihood.

## 5 Results

### 5.1 Quantification of onset age inconsistencies in CanPath data

We quantified onset age inconsistencies in CanPath data by computing the difference between self-reported onset ages at enrollment and follow-up for each condition. Figure 1 shows the distribution of these differences across 55 onset age variables in CanPath data, ordered by variance. A non-zero difference indicates an inconsistency, with larger absolute values suggesting greater inconsistency. As the distributions show no consistent unidirectional bias toward earlier or later reporting at follow-up, this validates our assumption in Matrix completion in Section 3.1. Variance differs substantially across conditions, with the age differences for some conditions (e.g., arthritis, breast cancer) showing notably higher variance than others.

**Figure 1.**
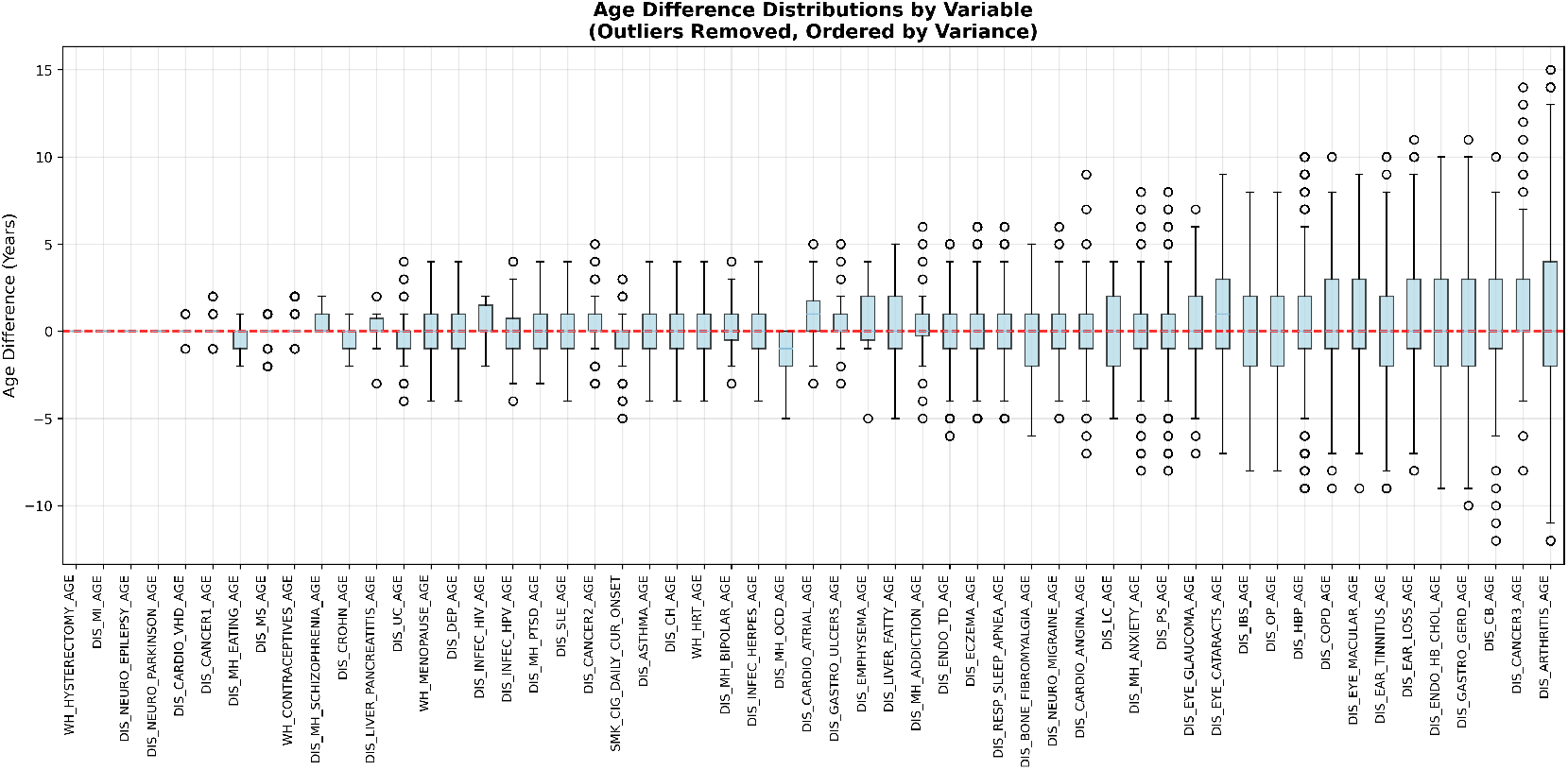
Box plots of enrollment and follow-up age difference distributions for onset age variables in CanPath data, with outliers removed and ordered by variance. Descriptions of the onset age variables can be found in Appendix A.

### 5.2 Reliability score-based stratification

#### 5.2.1 Uniform reliability score distribution

Figure 2(a) shows the distribution of reliability scores across all participants after normalization. The distribution is approximately uniform over [0,1] with mean 0.501 and median 0.499. This uniform distribution enables effective stratification of participants for our later experiments, as the absence of peaks or clustering at particular values ensures balanced cohort sizes for any subset of participants in task-specific evaluations.

**Figure 2.**
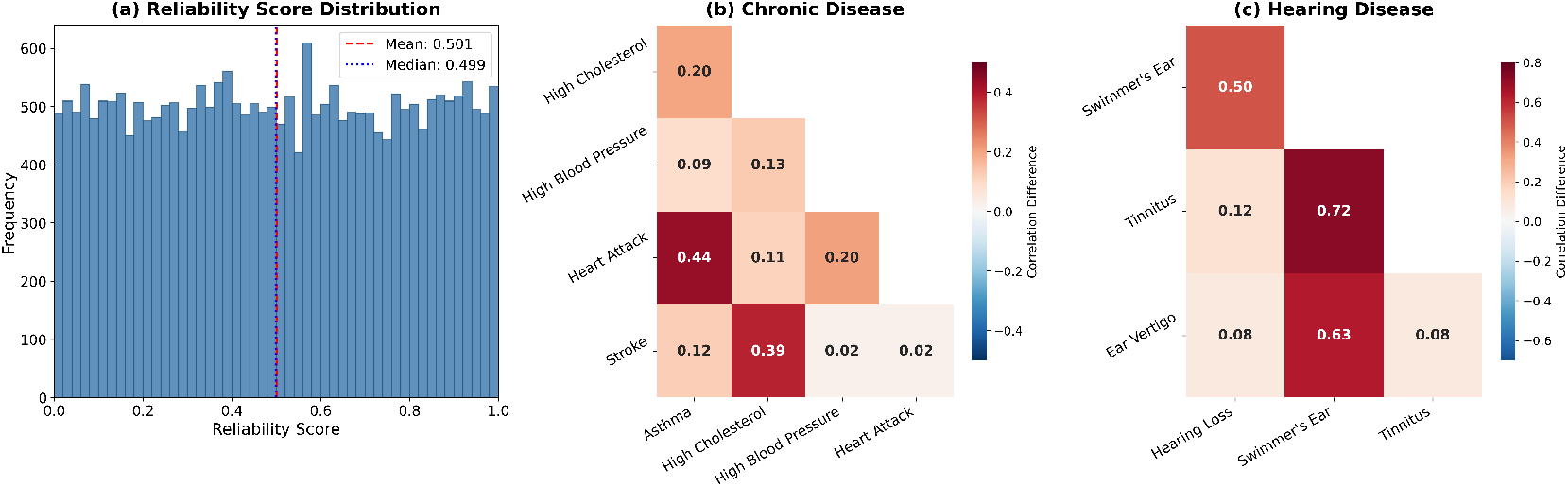
Reliability score distribution and onset age correlation differences between cohorts. (a) Distribution of reliability scores used to split participants into high- and low-reliability cohorts at the median. (b-c) Difference in correlations (high-reliability minus low-reliability) for pairs of onset ages in (b) chronic diseases and (c) hearing conditions. Positive values (red) indicate stronger correlations in the high-reliability cohort.

#### 5.2.2 Stronger correlations and more coherent disease clustering in high-reliability cohorts

Figures 2(b) and (c) show heatmaps where each cell shows the difference in correlation (high-reliability cohort minus low-reliability cohort) for pairs of onset ages in (b) chronic diseases (*co*_11_) and (c) hearing conditions (*co*_12_), respectively, where positive values indicate stronger correlations in the high-reliability cohort. Across both disease groups, correlations are consistently stronger in the high-reliability cohort, with differences ranging from 0.02 to 0.72. This indicates the high-reliability cohorts using reliability-based stratification recover stronger signals that better reflect underlying disease relationships.

Figure 3 presents disease onset age correlation networks for each cohort (*co*_13_), with communities identified using the Louvain algorithm [Blondel et al., 2008] with resolution parameter set to 1. We classified diseases into 12 categories based on the medical system (e.g., Cardiovascular, Respiratory, Gastrointestinal/Hepatic, Mental Health) using keyword matching on variable names. Full details can be found in Appendix C. While both cohorts yield five clusters across 45 onset age variables, the high-reliability network exhibits markedly greater biological coherence: The average proportion of diseases belonging to the dominant disease category within each cluster increases from 30.9% to 43.8%, while cluster entropy decreases from 2.23 to 1.86^6^, indicating that diseases are more consistently grouped with biologically related conditions in the high-reliability cohort.

**Figure 3.**
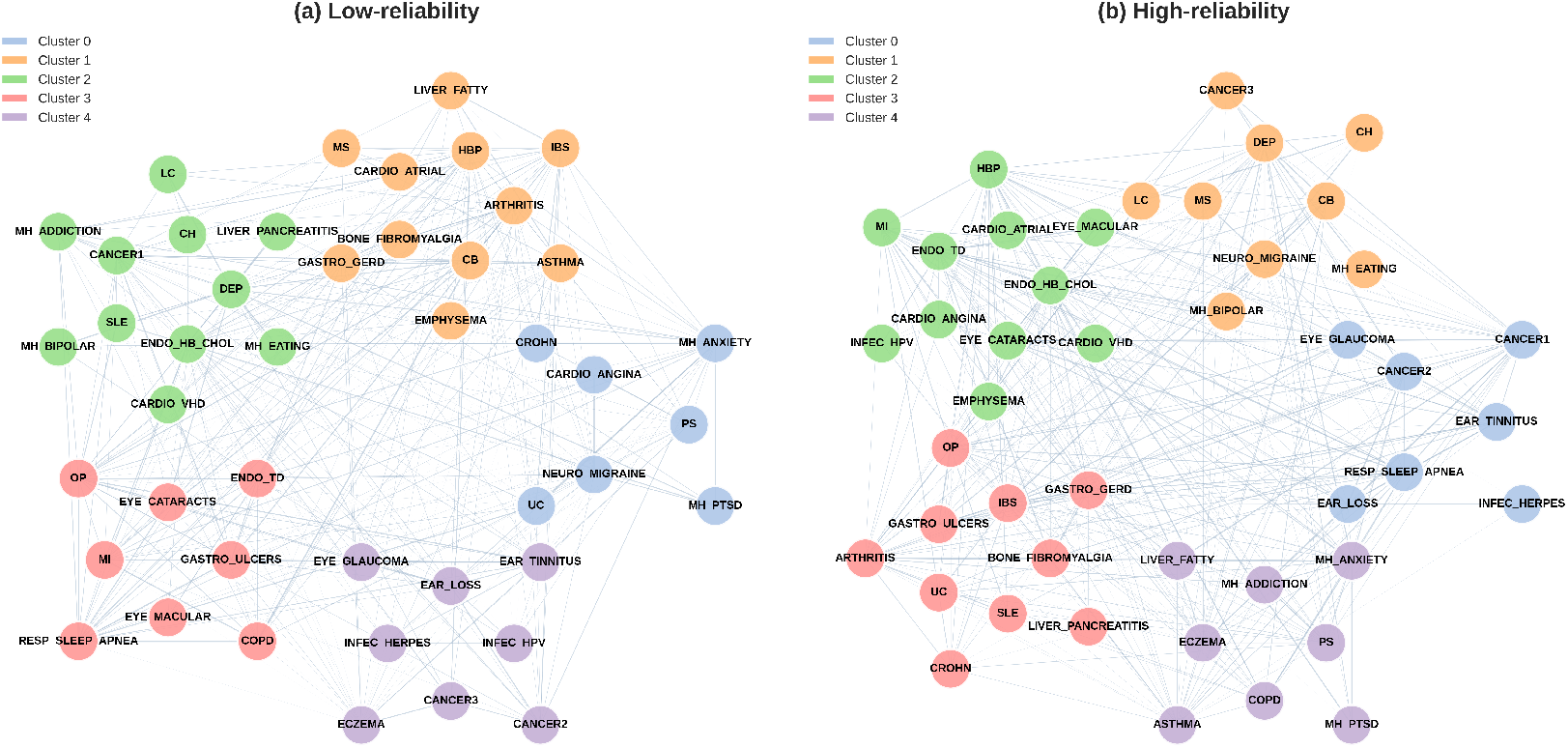
Disease onset age correlation networks stratified by reliability scores. Cluster networks showing communities of diseases with correlated ages of onset for (a) low-reliability and (b) high-reliability cohorts. Each node represents a disease, and edges indicate positive correlations between onset ages. Nodes are colored by cluster membership identified using the Louvain community detection algorithm. The average proportion of diseases in the dominant category within each cluster is 30.9% (low-reliability) and 43.8% (high-reliability), with cluster entropy of 2.23 (low-reliability) and 1.86 (high-reliability).

The improved clustering is particularly evident when examining the variables in specific clusters. In cluster 3 of the high-reliability network, gastrointestinal conditions (Crohn’s disease (CROHN), ulcerative colitis (UC), irritable bowel syndrome (IBS), persistent acid reflux (GASTRA_GERD), stomach (or duodenal) ulcers (GASTRA_ULCERS), and pancreatitis (LIVER_PANCREATITIS)) cluster together (60% gastrointestinal), whereas in the low-reliability network these conditions are scattered across multiple clusters. Similarly, in cluster 2 of the high-reliability network, cardiovascular and metabolic conditions (high blood pressure (HBP), high cholesterol (ENDO_HB_CHOL), heart attack (MI), angina (CARDIO_ANGINA), atrial fibrillation (CARDIO_ATRIAL)) form a cohesive cluster in the high-reliability network but are more diffusely distributed in the low-reliability network. These findings demonstrate that the high-reliability cohort significantly strengthens pairwise correlations and reveals clearer disease community structure.

#### 5.2.3 Improved predictive performance in high-reliability cohorts

Table 1 summarizes cross-validation performance for classification (*cl*_11_–*cl*_13_) and regression (*re*_11_–*re*_13_) tasks across cohorts. For each task, we selected the best model based on F1 score (classification) or RMSE (regression) on the high-reliability cohort. The tie-breaking rules can be found in Appendix E. The full results can be found in Appendix F.

**Table 1:**
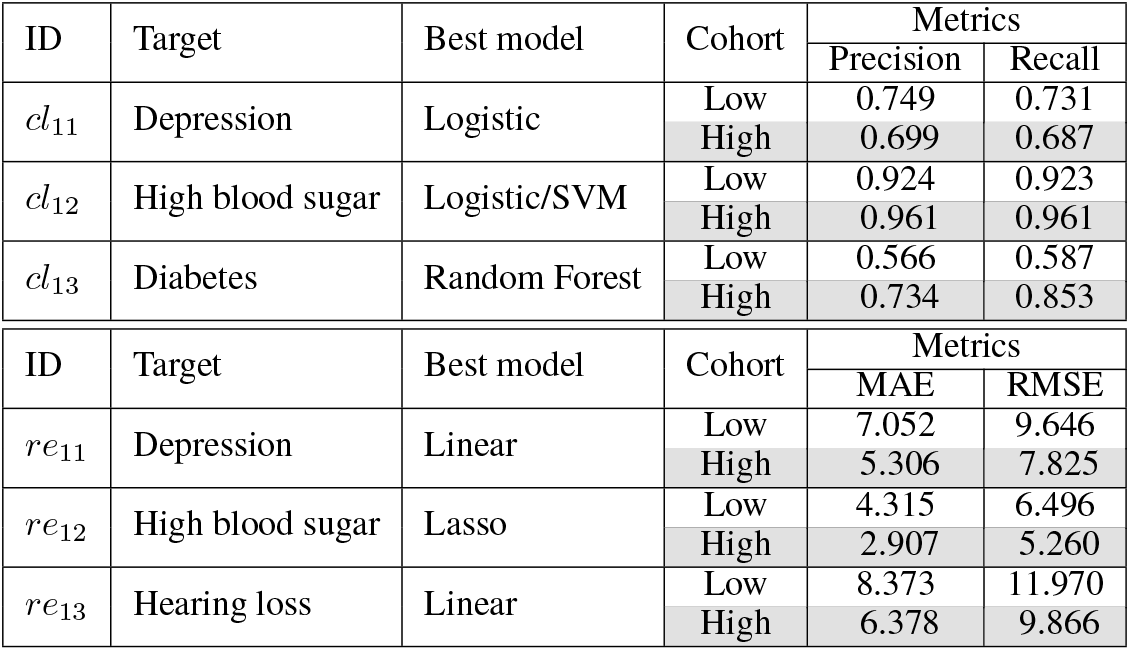
Cross-validation predictive performance on low-versus high-reliability cohorts for classification and regression tasks. Cohorts are divided by median reliability score. The best model is selected based on F1 score (classification) or RMSE (regression) on the high-reliability cohort. High-reliability cohort rows are highlighted in gray.

For regression tasks, the high-reliability cohort consistently yields substantially lower prediction errors, with MAE improvements ranging from 1.41 to 2.00 years and RMSE improvements from 1.24 to 2.10 years. Classification results are more mixed. For high blood sugar and diabetes prediction, the high-reliability cohort shows clear improvements in both precision and recall. However, for depression prediction, the low-reliability cohort unexpectedly outperforms the high-reliability cohort (0.749 vs. 0.699 in precision, and 0.731 vs. 0.687 in recall). This observation suggests that, for mental health variables, response variability may follow different patterns from other variables, highlighting the need for caution when applying reliability-based stratification in this setting.

### 5.3 Bayesian adjustment

#### 5.3.1 Strengthened correlations between biologically related conditions

Table 2 shows correlations computed using enrollment variables, follow-up variables, and Bayesian-adjusted values. Across all pairs, the correlation for Bayesian-adjusted values exceeds both the enrollment variables correlation and the follow-up variables. This indicates our Bayesian adjustment method recovers stronger associations that better reflect the underlying biological relationships.

**Table 2:**
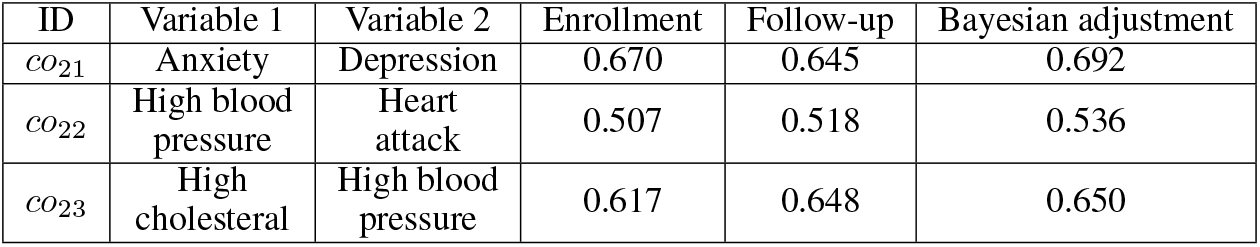
Pearson correlation coefficients between onset ages of variable 1 and 2 using enrollment variables, follow-up variables, and Bayesian-adjusted values.

#### 5.3.2 Consistent improvements with compounding benefits for multiple variables

Table 3 shows cross-validation performance for classification and regression tasks. We selected the best model based on F1 score for classification or RMSE for regression with Bayesian adjustment applied. The tie-breaking rules can be found in Appendix E. The full results can be found in Appendix G.

**Table 3:**
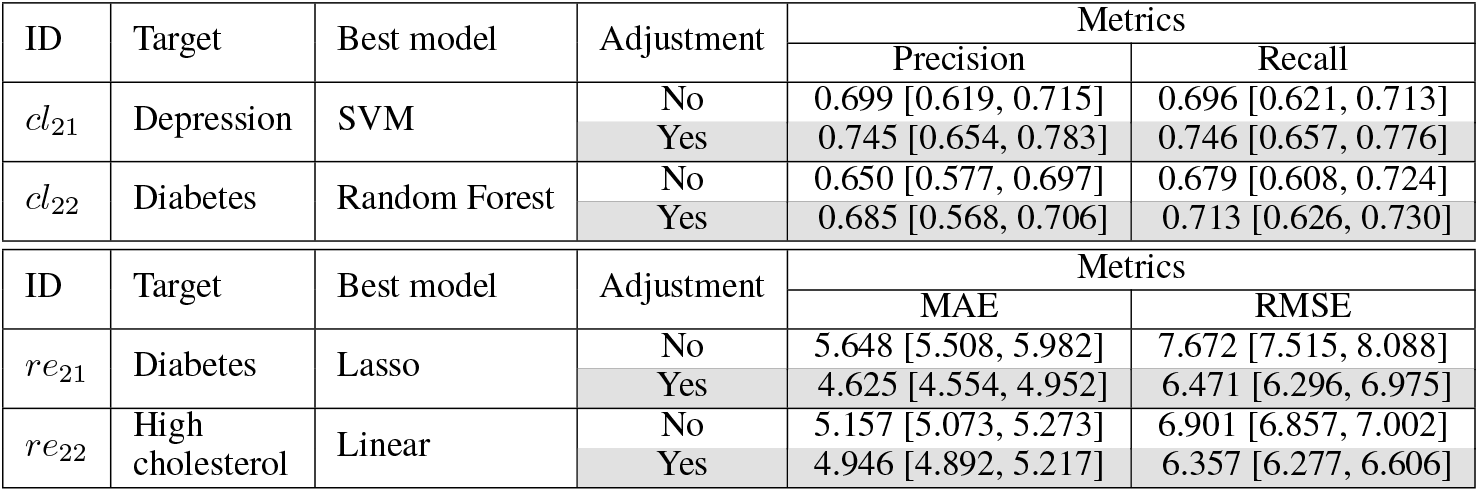
Cross-validation predictive performance without and with Bayesian adjustment on classification and regression tasks. 95% confidence intervals are shown in brackets. The best model is selected based on F1 score (classification) or RMSE (regression) after Bayesian adjustment. Results with Bayesian adjustment applied are highlighted in gray.

| ID | Target | Best model | Adjustment | Metrics |  |
| --- | --- | --- | --- | --- | --- |
|  |  |  |  | Precision | Recall |
| $cl_{21}$ | Depression | SVM | No | 0.699 [0.619, 0.715] | 0.696 [0.621, 0.713] |
|  |  |  | Yes | 0.745 [0.654, 0.783] | 0.746 [0.657, 0.776] |
| $cl_{22}$ | Diabetes | Random Forest | No | 0.650 [0.577, 0.697] | 0.679 [0.608, 0.724] |
|  |  |  | Yes | 0.685 [0.568, 0.706] | 0.713 [0.626, 0.730] |
| ID | Target | Best model | Adjustment | Metrics |  |
|  |  |  |  | MAE | RMSE |
| $re_{21}$ | Diabetes | Lasso | No | 5.648 [5.508, 5.982] | 7.672 [7.515, 8.088] |
|  |  |  | Yes | 4.625 [4.554, 4.952] | 6.471 [6.296, 6.975] |
| $re_{22}$ | High cholesterol | Linear | No | 5.157 [5.073, 5.273] | 6.901 [6.857, 7.002] |
|  |  |  | Yes | 4.946 [4.892, 5.217] | 6.357 [6.277, 6.606] |

Across all tasks, Bayesian adjustment consistently improves predictive performance. A notable improvement occurs in diabetes onset age prediction (*re*1), where MAE decreases by 18% (from 5.648 to 4.625), and RMSE decreases by 16% (from 7.672 to 6.471). This task involves the adjustment of two predictor variables (high blood pressure and high cholesterol onset ages), suggesting that benefits compound when multiple inconsistent variables are adjusted simultaneously. The 95% CIs widen modestly after Bayesian adjustment, with precision CIs widening by 0.026, recall by 0.008, MAE by 0.025 years, and RMSE by 0.145 years on average. This indicates that the uncertainty introduced by our Bayesian adjustment method is modest relative to the gains in point estimates.

## 6 Conclusion and discussion

We presented two methods for handling onset age inconsistencies for longitudinal healthcare survey data. First, we constructed participant-level reliability scores by aggregating inconsistency magnitudes across onset age variables. By stratifying participants using the scores, we found that participants in the high-reliability cohorts in general demonstrate stronger positive correlations among biologically related variables, more coherent and interpretable disease communities in clustering networks, and better predictive performance in various classification and regression tasks. Second, we proposed a Bayesian adjustment method that models enrollment and follow-up onset ages as two noisy measurements of a latent true onset age. Across multiple case studies, our method increased correlation estimates for biologically associated onset-age pairs and improved downstream predictive performance for both classification and regression.

The two methods address different needs for healthcare practitioners and can be viewed as complementary. Reliability-based stratification is more appropriate when (1) the dataset is sufficiently large that deprioritizing or excluding participants in low-reliability cohorts does not impact the learning of tasks; and (2) ease of deployment is the priority, since the scores can be computed once and reused across analyses. In contrast, Bayesian adjustment is more appropriate when (1) the sample size is limited; (2) the researchers want to propagate uncertainty from the data into subsequent inference; and (3) tasks involve mental-health variables, where the response variability may be different from other variables, making variable-by-variable adjustment preferable to participant-level exclusion.

In the future, we plan to explore the following directions. First, our reliability score focuses on onset age inconsistencies and does not capture other forms of healthcare longitudinal inconsistencies, such as changes in disease status from “yes” to “no” across survey waves (see Appendix H for details); extending reliability scoring to incorporate multiple inconsistency types is an important next step. Second, reliability-based stratification may introduce selection bias by excluding certain subpopulations. This suggests the need for reliability-aware analyses. Third, our Bayesian adjustment framework is designed for two time points (enrollment and follow-up); future work will extend it to multiple survey waves to enable adjustment using a more flexible number of time points.

## Data Availability

CanPath data are available to researchers through a controlled access process via the CanPath Access Portal (https://portal.canpath.ca).

## A Descriptions of 55 onset age variables collected at enrollment and follow-up in CanPath data

**Table 4:**
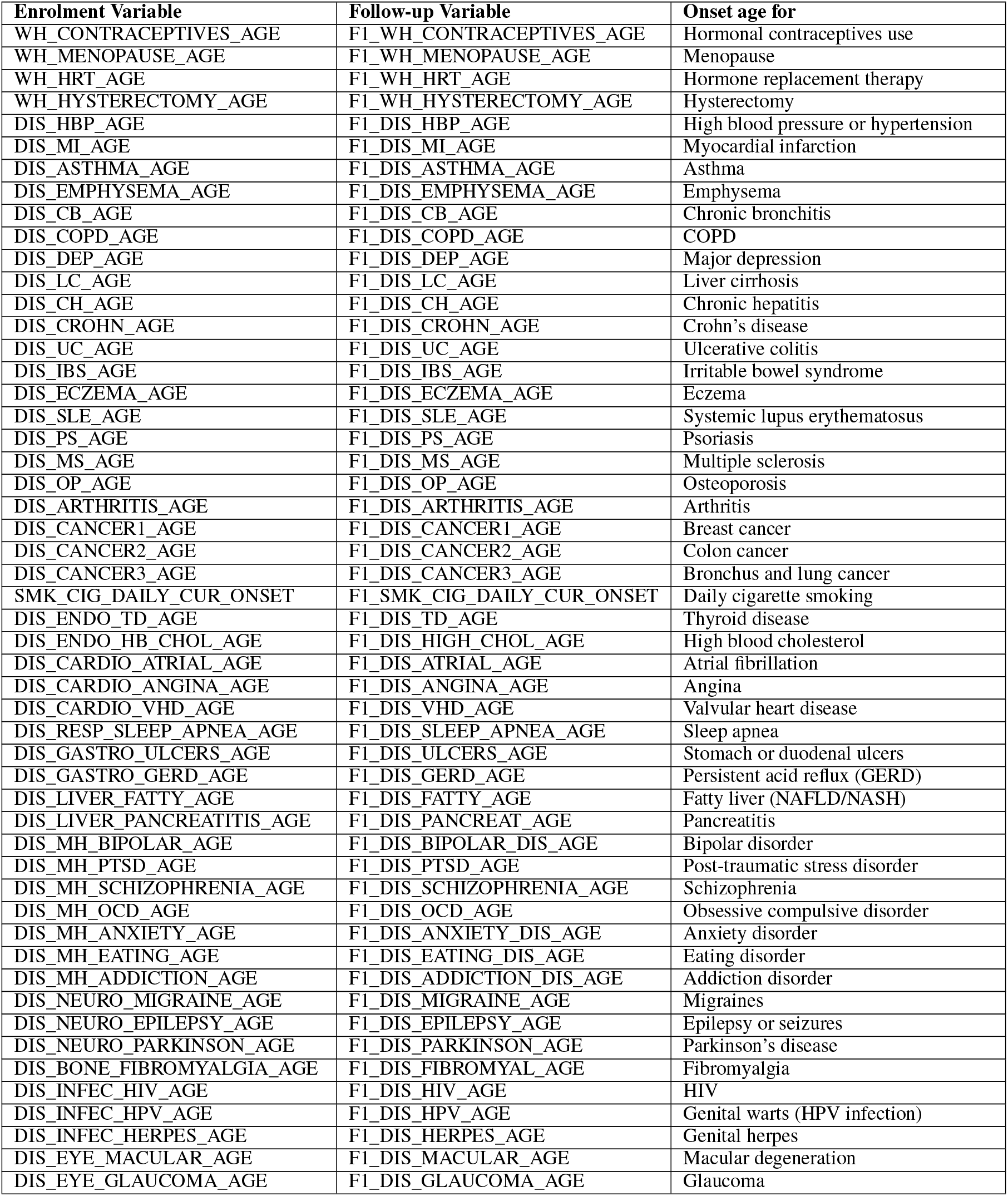
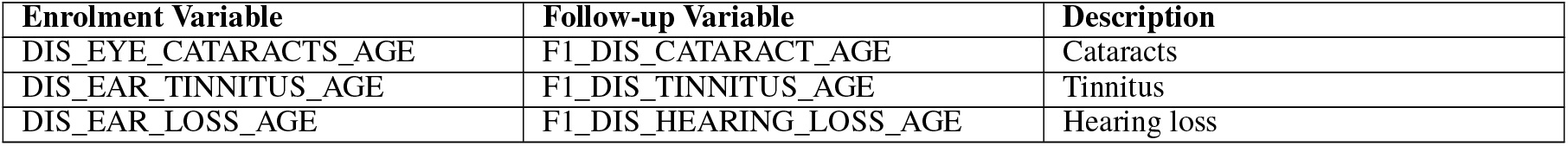
Full names and condition descriptions for the onset age pairs at enrollment and at follow-up.

## B Derivation of the posterior distribution

We assume a diffuse (non-informative) Normal prior on the latent true value:

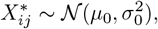

where *µ*_0_ represents our prior mean belief about the true onset age and 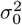 represents our prior uncertainty. In the absence of strong prior knowledge about the true onset age, we use a diffuse prior by letting 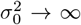. By Bayes’ theorem and the conditional independence of 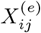 and 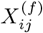 given 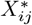, the posterior is:

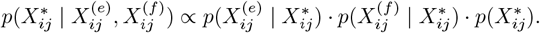

The posterior distribution is Normal due to conjugacy: when both the likelihood and prior are Normal distributions, the posterior is also Normal. Taking the logarithm:

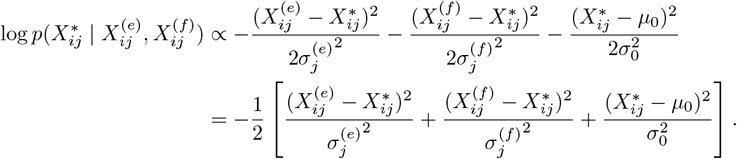

As 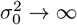, the prior term vanishes, leaving:

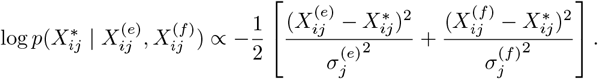

Expanding the squared terms:

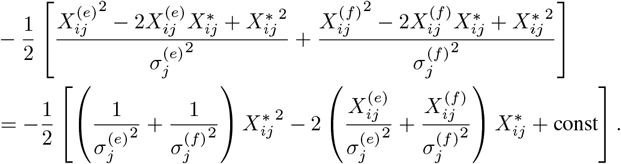

Completing the square, we obtain a Normal distribution with:

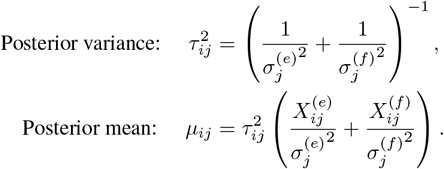

Therefore, the posterior distribution is:

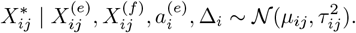

## C Classification of diseases

**Table 5:**
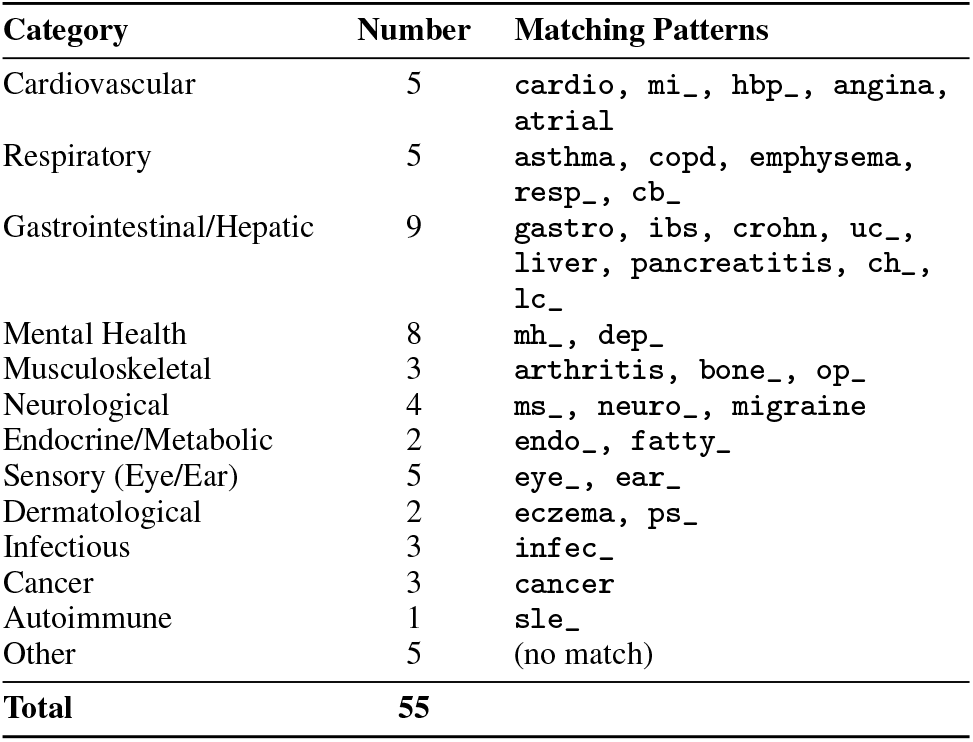
Disease categorization rules for 55 onset age variables. Patterns are matched case-insensitively against variable names.

## D Coherence metrics

Suppose *C* is the number of medical categories in the cluster, *n*_*i*_ is the number of diseases in category *i*, and *N* is the total number of diseases in the cluster. Entropy is

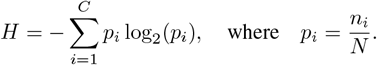

Dominant category percentage is:

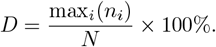

## E Tie-breaking rules for determining the best model

When selecting the best model for each task, we apply the following tie-breaking rules:

### Reliability score-based stratification

- **Classification tasks:** Models are ranked by F1 score on the high-reliability cohort. If multiple models achieve the same F1 score on the high-reliability cohort, we compare their F1 scores on the low-reliability cohort and select the model with the higher F1 score.
- **Regression tasks:** Models are ranked by RMSE on the high-reliability cohort (lower is better). If multiple models achieve the same RMSE on the high-reliability cohort, we compare their MAE values on the high-reliability cohort and select the model with the lower MAE.

### Bayesian adjustment

- **Classification tasks:** Models are ranked by F1 score after Bayesian adjustment is applied. We do not observe ties for this category.
- **Regression tasks:** Models are ranked by RMSE after Bayesian adjustment is applied (lower is better). If multiple models achieve the same RMSE, we compare their MAE values after Bayesian adjustment and select the model with the lower MAE.

## F Full predictive results for reliability score-based stratification

**Table 6:** Cross-validation classification performance across over *cl*_11_, *cl*_12_ and *cl*_13_ for five models across low- and high-reliability cohorts (divided by median reliability score). Performance metrics include precision and recall. F1 scores are used for determining the best model. High-reliability cohort rows are highlighted in gray, and the best models are denoted with (*).

| ID | Target | Model | Cohort | Metrics |  |  |
| --- | --- | --- | --- | --- | --- | --- |
|  |  |  |  | Precision | Recall | F1 |
| $cl_{11}$ | Depression | Logistic (*) | Low | 0.749 | 0.731 | - |
|  |  |  | High | 0.699 | 0.687 | 0.673 |
|  |  | Random Forest | Low | 0.723 | 0.719 | - |
|  |  |  | High | 0.656 | 0.655 | 0.647 |
|  |  | Decision Tree | Low | 0.715 | 0.706 | - |
|  |  |  | High | 0.648 | 0.645 | 0.632 |
|  |  | Gradient Boosting | Low | 0.741 | 0.729 | - |
|  |  |  | High | 0.672 | 0.666 | 0.654 |
|  |  | SVM | Low | 0.748 | 0.723 | - |
|  |  |  | High | 0.721 | 0.689 | 0.666 |
| $cl_{12}$ | High blood sugar | Logistic (*) | Low | 0.924 | 0.923 | 0.917 |
|  |  |  | High | 0.961 | 0.961 | 0.958 |
|  |  | Random Forest | Low | 0.909 | 0.912 | 0.917 |
|  |  |  | High | 0.955 | 0.957 | 0.954 |
|  |  | Decision Tree | Low | 0.850 | 0.843 | - |
|  |  |  | High | 0.931 | 0.930 | 0.930 |
|  |  | Gradient Boosting | Low | 0.922 | 0.921 | - |
|  |  |  | High | 0.954 | 0.956 | 0.952 |
|  |  | SVM (*) | Low | 0.924 | 0.923 | 0.917 |
|  |  |  | High | 0.961 | 0.961 | 0.958 |
| $cl_{13}$ | Diabetes | Logistic | Low | 0.749 | 0.707 | - |
|  |  |  | High | 0.728 | 0.820 | 0.769 |
|  |  | Random Forest (*) | Low | 0.566 | 0.587 | 0.552 |
|  |  |  | High | 0.734 | 0.853 | 0.787 |
|  |  | Decision Tree | Low | 0.790 | 0.700 | - |
|  |  |  | High | 0.706 | 0.673 | 0.684 |
|  |  | Gradient Boosting | Low | 0.585 | 0.580 | - |
|  |  |  | High | 0.712 | 0.707 | 0.703 |
|  |  | SVM | Low | 0.457 | 0.587 | 0.500 |
|  |  |  | High | 0.734 | 0.853 | 0.787 |

**Table 7:** Cross-validation regression performance over task *re*_11_, *re*_12_ and *re*_13_ for five models across low- and high-reliability cohorts (divided by median reliability score). Performance metrics include MAE and RMSE. High-reliability cohort rows are highlighted in gray, and the best models are denoted with (*).

| ID | Target | Model | Cohort | Metrics |  |
| --- | --- | --- | --- | --- | --- |
|  |  |  |  | MAE | RMSE |
| $re_{11}$ | Depression | Linear(*) | Low | 7.052 | 9.646 |
|  |  |  | High | 5.306 | 7.825 |
|  |  | Lasso | Low | 7.345 | 9.750 |
|  |  |  | High | 5.708 | 7.919 |
|  |  | Ridge | Low | 7.059 | 9.646 |
|  |  |  | High | 5.320 | 7.825 |
|  |  | Random Forest | Low | 8.075 | 11.127 |
|  |  |  | High | 6.105 | 9.422 |
|  |  | SVM | Low | 6.570 | 9.795 |
|  |  |  | High | 5.001 | 8.148 |
| $re_{12}$ | High blood sugar | Linear | Low | 4.538 | 6.995 |
|  |  |  | High | 3.463 | 5.625 |
|  |  | Lasso (*) | Low | 4.315 | 6.496 |
|  |  |  | High | 2.907 | 5.260 |
|  |  | Ridge | Low | 4.547 | 6.950 |
|  |  |  | High | 3.438 | 5.590 |
|  |  | Random Forest | Low | 4.502 | 7.434 |
|  |  |  | High | 3.258 | 6.187 |
|  |  | SVM | Low | 6.961 | 9.300 |
|  |  |  | High | 6.802 | 10.416 |
| $re_{13}$ | Hearing loss | Linear (*) | Low | 8.367 | 11.970 |
|  |  |  | High | 6.378 | 9.866 |
|  |  | Lasso | Low | 8.688 | 12.018 |
|  |  |  | High | 6.710 | 9.913 |
|  |  | Ridge | Low | 8.373 | 11.970 |
|  |  |  | High | 6.385 | 9.866 |
|  |  | Random Forest | Low | 8.502 | 12.614 |
|  |  |  | High | 6.744 | 10.832 |
|  |  | SVM | Low | 7.291 | 11.784 |
|  |  |  | High | 5.958 | 10.357 |

## G Full predictive results for Bayesian adjustment

**Table 8:** Cross-validation classification performance over task *cl*_21_ and *cl*_22_ without and with Bayesian adjustment using five machine learning models. Metrics include precision, recall, and 95% confidence intervals in brackets. F1 scores are used for determining the best model. Results with Bayesian adjustment applied are highlighted in gray, and the best models are denoted with (*).

| ID | Target | Model | Adjustment | Metrics |  |  |
| --- | --- | --- | --- | --- | --- | --- |
|  |  |  |  | Precision | Recall | F1 |
| $cl_{21}$ | Depression | Logistic | No | 0.689 [0.642, 0.709] | 0.687 [0.645, 0.704] | - |
|  |  |  | Yes | 0.730 [0.692, 0.761] | 0.731 [0.687, 0.761] | 0.728 |
|  |  | Random Forest | No | 0.593 [0.497, 0.671] | 0.591 [0.501, 0.666] | - |
|  |  |  | Yes | 0.655 [0.501, 0.692] | 0.642 [0.493, 0.687] | 0.643 |
|  |  | Decision Tree | No | 0.570 [0.459, 0.665] | 0.564 [0.454, 0.657] | - |
|  |  |  | Yes | 0.582 [0.458, 0.672] | 0.567 [0.448, 0.672] | 0.569 |
|  |  | Gradient Boosting | No | 0.660 [0.514, 0.698] | 0.663 [0.519, 0.699] | - |
|  |  |  | Yes | 0.679 [0.524, 0.722] | 0.672 [0.522, 0.716] | 0.673 |
|  |  | SVM (*) | No | 0.699 [0.619, 0.715] | 0.696 [0.621, 0.713] | - |
|  |  |  | Yes | 0.745 [0.654, 0.783] | 0.746 [0.657, 0.776] | 0.744 |
| $cl_{22}$ | Diabetes | Logistic | No | 0.519 [0.517, 0.774] | 0.720 [0.701, 0.736] | - |
|  |  |  | Yes | 0.521 [0.521, 0.804] | 0.722 [0.713, 0.739] | 0.605 |
|  |  | Random Forest (*) | No | 0.650 [0.577, 0.697] | 0.679 [0.608, 0.724] | - |
|  |  |  | Yes | 0.685 [0.568, 0.706] | 0.713 [0.626, 0.730] | 0.692 |
|  |  | Decision Tree | No | 0.629 [0.562, 0.691] | 0.625 [0.541, 0.687] | - |
|  |  |  | Yes | 0.658 [0.552, 0.593] | 0.661 [0.548, 0.696] | 0.659 |
|  |  | Gradient Boosting | No | 0.658 [0.580, 0.711] | 0.699 [0.622, 0.734] | - |
|  |  |  | Yes | 0.645 [0.568, 0.713] | 0.696 [0.626, 0.739] | 0.655 |
|  |  | SVM | No | 0.699 [0.619, 0.715] | 0.696 [0.621, 0.713] | - |
|  |  |  | Yes | 0.745 [0.654, 0.783] | 0.746 [0.657, 0.776] | 0.605 |

**Table 9:**
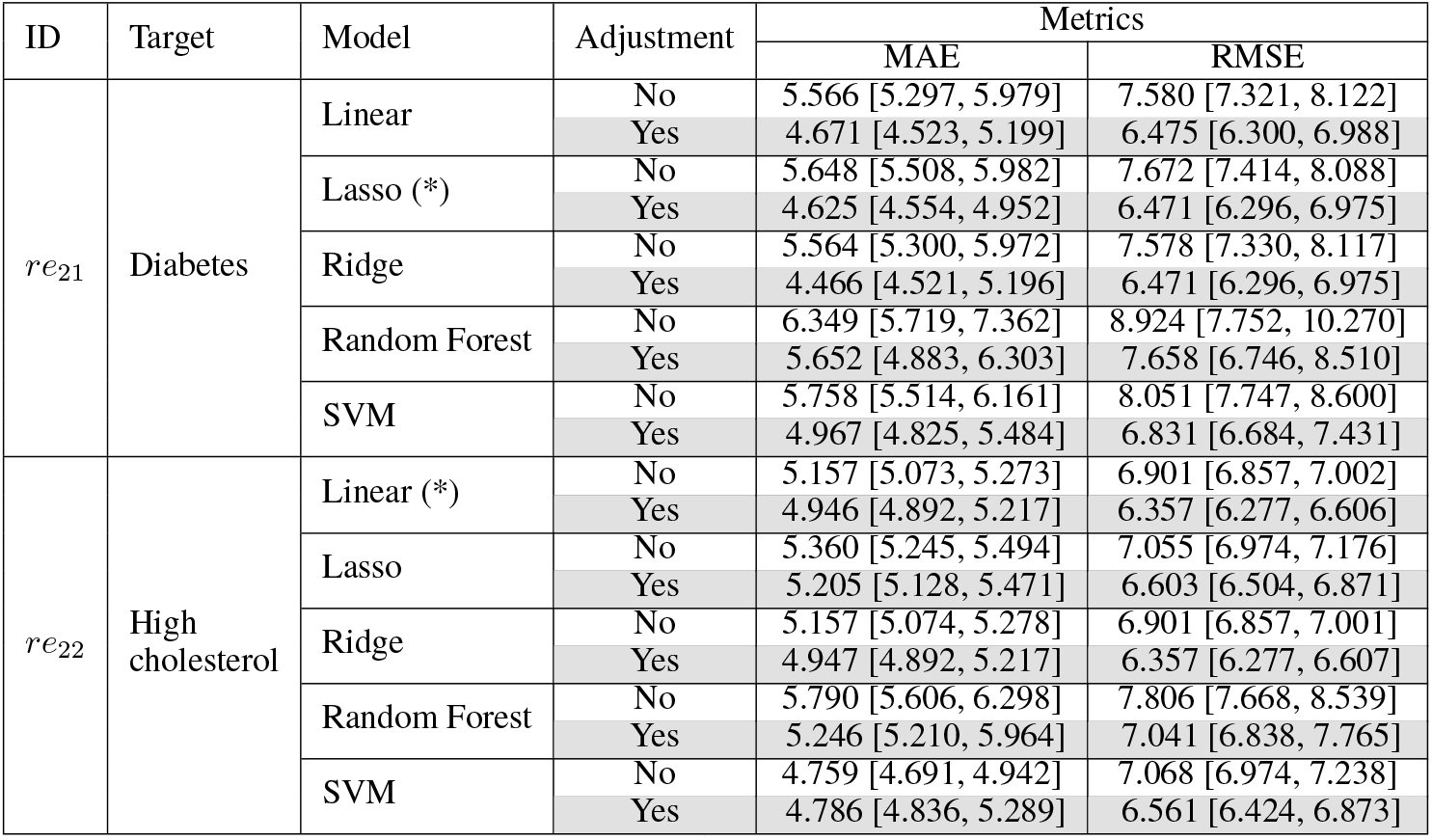
Cross-validation regression performance over task *re*_21_ and *re*_22_ without and with Bayesian adjustment using five machine learning models. Metrics include MAE and RMSE with 95% confidence intervals in brackets. Results with Bayesian adjustment applied are highlighted in gray, and the best models are denoted with (*).

## H Summary of Inconsistency Types in CanPath data

The following inconsistency types were identified across 97,408 participants with both enrollment and follow-up data. Percentages indicate the proportion of participants exhibiting at least one inconsistency of each type.

### Onset age inconsistency (57.1% of participants)

This inconsistency occurs when the age at which a participant first reported being diagnosed with a condition differs between enrollment and follow-up surveys. For example, a participant may report being diagnosed with diabetes at age 45 during enrollment, but report age 50 at follow-up. Since the age of first diagnosis is a fixed historical event, any difference between timepoints is considered inconsistent.

### Ever-had inconsistency (55.1% of participants)

This inconsistency occurs when a participant reports “yes” to having a condition at enrollment, but “no” at follow-up (i.e., a 1 → 0 transition). For example, a participant may report “yes” to ever having asthma at enrollment, but “no” at follow-up. Such transitions are inconsistent because one cannot “un-have” a previously reported condition. Note that 0 → 1 transitions are acceptable, as conditions can develop over time.

### Calculated age vs date inconsistency (0.24% of participants)

This inconsistency occurs when the difference in calculated age between enrollment and follow-up does not match the time gap between the two questionnaire completion dates. For example, if questionnaires are completed 5 years apart, but reported ages only differ by 2 years, this is flagged as inconsistent. A tolerance of *±*2 years is applied to account for integer rounding in age reporting.

## Footnotes

2 Note that 57.1% refers to participants who reported different onset ages for at least one condition between enrollment and follow-up, given that enrollment and follow-up response are both not missing.

3 A more complex design could separately track under-estimation and over-estimation patterns using two reliability scores, one for each direction of bias.

4 While other methods such as Multivariate Imputation by Chained Equations (MICE) [Van Buuren and Groothuis-Oudshoorn, 2011] may suit certain datasets, we recommend SoftImpute as a starting point. SoftImpute captures low-rank linear structure through matrix factorization, making it more robust to noise and less sensitive to variable ordering than iterative regression methods like MICE, which can accumulate errors when dependencies between variables are weak or when missingness is high.

5 This is different from Section 4.2 where we used both the onset ages during enrollment and follow-up.

6 Full definitions of the coherence metrics are provided in Appendix D.

## Notes

### Competing Interest Statement

The authors have declared no competing interest.

### Funding Statement

This study did not receive any external funding.

### Author Declarations

Ethics committee/IRB of the University of British Columbia gave ethical approval for this work.

